# Adjusting Power Calculations and Trial Sample Size for Treatment Resistance

**DOI:** 10.1101/2025.10.14.25337997

**Authors:** Burcu Tepekule

## Abstract

Statistical calculations for clinical trials traditionally assume that if a treatment fails, it fails for mechanistic reasons—the drug itself is ineffective. However, patients may be treatment-resistant, rendering them unable to benefit from an otherwise effective treatment. This creates an identifiability problem: a null hypothesis that we fail to reject can indicate either an ineffective treatment, or an effective treatment tested in a population dominated by treatment-resistant subjects. However, the strategy to administer the drug should be different for these cases. Here we present a simple way to adjust the sample size of a randomized controlled trial to account for the anticipated level of treatment resistance to reach a certain statistical power. We show that the resistant-adjusted population size exponentially increases with the anticipated resistance prevalence, whereas power decreases almost linearly for a given population size as the resistance prevalence increases.

## Introduction

Treatment resistance can come in different flavors. In cancer immunotherapy, only a subset of patients harbor tumors that are sensitive to checkpoint inhibitors, with the remainder being mechanistically unable to respond (*1*). In HIV prevention trials, broadly neutralizing antibodies can only protect against viral strains with sufficient in vitro sensitivity to the antibody (*2*). Antibiotic resistance renders entire classes of drugs ineffective against resistant bacterial strains (*3*). While these scenarios differ in their biological mechanisms, they share a common statistical challenge: an effective treatment appears ineffective when tested in populations where most individuals cannot respond. Here, we suggest a simple adjustment to the standard Welch’s two-proportion sample size calculation (*4*) by reducing the expected treatment response rate to account for the prevalence of resistant individuals. Specifically, for a two-arm randomized trial with binary outcomes comparing a control group with response rate *p*_*C*_ to a treatment group with resistance-free response rate *p*_*T*_, we adjust the treatment rate as *p*_*T*_^*adj*^ = *p*_*T*_ − Δ × *p*_*R*_, where Δ = (*p*_*T*_ − *p*_*C*_) denotes the resistance-free effect size and *p*_*R*_ is the anticipated prevalence of treatment-resistant patients. This adjustment reduces the treatment effect by the fraction of patients who cannot mechanistically respond, providing a more realistic basis for power calculations. While we focus on binary outcomes, this principle extends naturally to continuous outcomes through analogous adjustments to expected mean differences.

## Results

We first demonstrate a case scenario where *p*_*C*_ = 0. 25, *p*_*T*_ = 0. 75, and *p*_*R*_ = 0. 5 (50% of the population is treatment-resistant). For each sample size *N* (sample size, or number of patients per group), we run 1000 simulations to calculate the empirical power—the proportion of simulated trials that correctly reject the null hypothesis at α = 0. 05. We then compare these empirical results to the theoretical sample size required to achieve 80% power, calculated using Welch’s two-proportion formula in both unadjusted (*N*^*u*^) and adjusted (*N*^*adj*^) forms (Fig. 1). We observe that without adjustment, the required sample size *N*^*u*^ = 13, whereas in reality, *N*^*adj*^ = 56 subjects per group are required to achieve 80% power at α = 0. 05.

**Figure 1.**
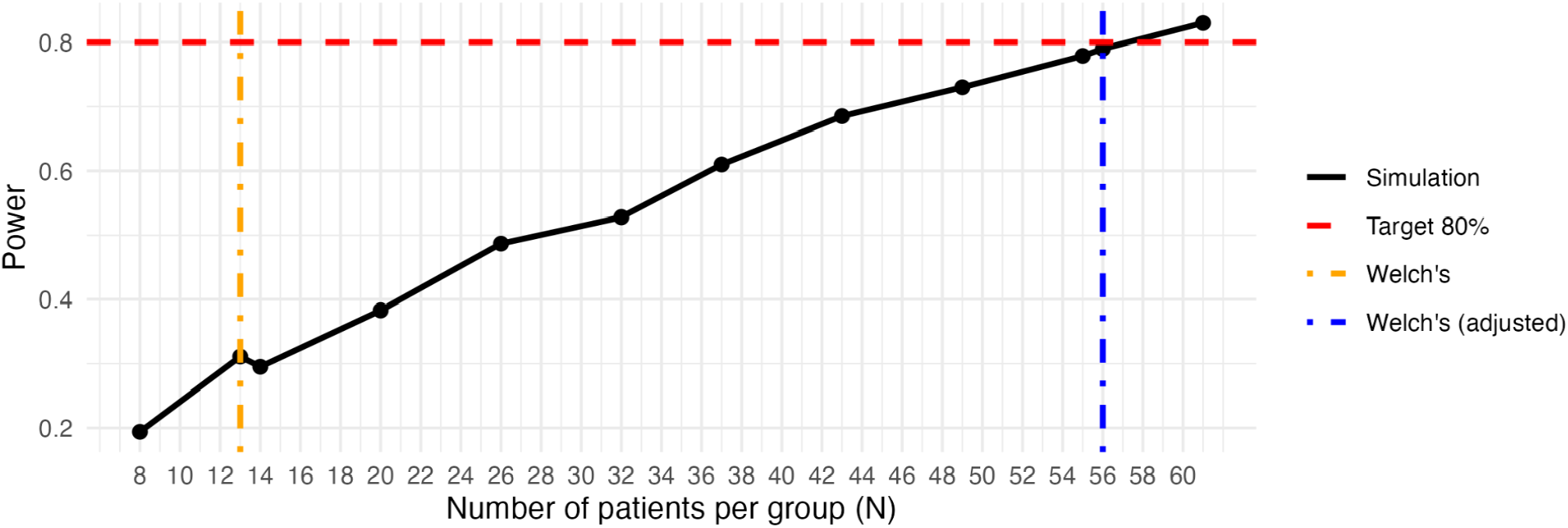
Empirical power curves demonstrating the need for resistance-adjusted sample size calculations. The black solid line shows empirical power from 1000 simulations at each sample size *N* . The orange dashed line indicates the Welch’s (unadjusted) two-proportion sample size calculation predicting *N*^*u*^ = 13 patients per group would achieve 80% power at α = 0. 05 (red horizontal line), assuming there is no treatment resistance in the population. With 50% treatment resistance, this sample size yields substantially lower empirical power, only ∼30% rather than 80%. The blue dashed line shows the adjusted Welch’s calculation correctly predicting *N*^*adj*^ = 56 patients per group to achieve 80% power—a 4.3-fold increase. The alignment between the adjusted theoretical calculation (blue line) and the empirical curve (black line) at 80% power validates the analytical adjustment.

We then investigate how adjusted sample size *N*^*adj*^ increases as a function of the prevalence of treatment-resistant patients *p*_*R*_. We show that this relationship is exponential (Fig. 2a, *p*_*R*_ ≤ 0. 9), which makes intuitive sense: as resistance prevalence approaches 100%, it becomes impossible to detect any difference between control and treatment groups, theoretically requiring infinite sample size. Conversely, as resistance prevalence *p*_*R*_ increases, the adjusted effect size Δ ^*adj*^ = *p*_*T*_ ^*adj*^ – *p*_*C*_ = (*p*_*T*_ − Δ × *p*_*R*_) – *p*_*C*_ = Δ × (1 − *p*_*R*_) shrinks linearly, causing an approximately linear decline in statistical power when sample size is held constant (Fig. 2b).

**Figure 2.**
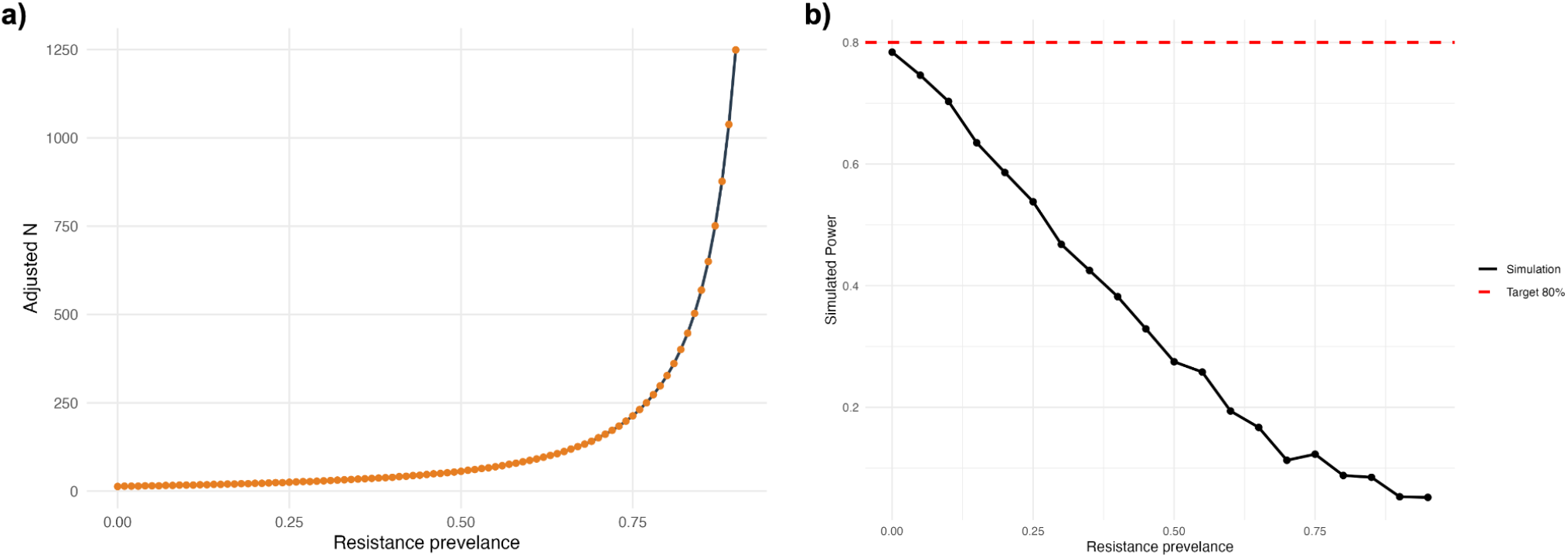
Relationship between resistance prevalence *p*_*R*_ and study design parameters. **a)** Adjusted sample size *N*^*adj*^ required per group to achieve 80% power at α = 0. 05 for *p*_*R*_ ≤ 0. 9. *N* ^*adj*^ increases exponentially with resistance prevalence *p*_*R*_, approaching infinity as *p* → 1. **b)** For a fixed sample size, empirical (simulated) power decreases approximately linearly with resistance prevalence.

## Discussion

This study demonstrates that treatment resistance has substantial implications for clinical trial design. Ignoring resistance when a large proportion of the patient population cannot mechanistically benefit from treatment leads to predictably underpowered studies. Critically, this creates a Type II error: failing to reject the null hypothesis and concluding a treatment is ineffective when it may actually work well in sensitive patients. Such false negatives can lead to premature abandonment of potentially useful therapies.

Most importantly, our results show that to maintain constant power at a target level (e.g., 80%), the required sample size *N*^*adj*^ must grow exponentially to compensate for this diminishing effect size (Fig. 2a). This exponential growth becomes particularly pronounced above 75% resistance, where sample size requirements increase dramatically and approach infinity as *p*_*R*_ → 1, at which no difference between treatment and control can be detected. This dramatic inflation in sample size requirements has critical implications for trial feasibility and resource allocation. At high resistance levels, trials may become prohibitively expensive and infeasible. At this point, one should reconsider whether a traditional randomized controlled trial remains the appropriate study design—at least for drugs that have demonstrated mechanistic efficacy *in vitro* and *in vivo* during preclinical studies. Alternative approaches, such as reallocating resources toward more *personalized* trials, might be more cost- and treatment-effective in the long run.

Our findings align with recent work showing similar sample size inflation when only a proportion of patients can respond to treatment. In cancer immunotherapy, Xu *et al*. (*5*) demonstrated that when only 20% of patients responded (versus 60%), required sample sizes increased 5.2-fold (from 52 to 269 patients). In HIV prevention trials, Corey *et al*. (*6*) found that the broadly neutralizing antibody VRC01 achieved 75% prevention efficacy against viruses with high *in vitro* sensitivity, but only 30% of circulating viruses met this threshold, explaining the lack of overall prevention efficacy. Without appropriate adjustment for this resistance, such trials risk discarding treatments that could benefit substantial subpopulations of patients.

The proposed adjustment is neither sophisticated nor hard to implement. However, it requires an estimate of resistance prevalence in the target population. Such estimates may be obtained from sources like surveillance data on resistant strains (if the new drug has a similar mechanism of action) or biomarker prevalence studies (for targeted therapies). However, when the tested drug is new with a novel mechanism of action that cannot be compared to existing drugs, resistance prevalence may be difficult to estimate, which is the point of the randomized clinical trials. Nevertheless, even without precise estimates, understanding how resistance-driven dilution of treatment effects leads to underpowered trials should give investigators pause before committing a Type II error and discarding a drug that has demonstrated mechanistic efficacy *in vitro* and *in vivo* animal studies.

## Limitations

Our approach relies on the normal approximation to the binomial distribution, which assumes moderate response rates. When treatment response rates approach boundary values (near 0 or 1), the variance *p*(1 − *p*) approaches zero and the normal approximation becomes unreliable. In such cases, exact methods (such as Fisher’s exact test) or alternative sample size calculations may be more appropriate.

## Data Availability

All data produced are available online at https://github.com/burcutepekule/adjusted_welch/

## Code Availability

All code (in R) is available at https://github.com/burcutepekule/adjusted_welch/

## Notes

### Competing Interest Statement

The authors have declared no competing interest.

### Funding Statement

This study did not receive any funding

